# Serology assessment of antibody response to SARS-CoV-2 in patients with COVID-19 by rapid IgM/IgG antibody test

**DOI:** 10.1101/2020.08.05.20168815

**Authors:** Yang De Marinis, Torgny Sunnerhagen, Pradeep Bompada, Anna Bläckberg, Runtao Yang, Joel Svensson, Ola Ekström, Karl-Fredrik Eriksson, Ola Hansson, Leif Groop, Isabel Gonçalves, Magnus Rasmussen

## Abstract

The coronavirus disease 2019 (COVID-19) pandemic has created a global health- and economic crisis. Lifting confinement restriction and resuming to normality depends greatly on COVID-19 immunity screening. Detection of antibodies to severe acute respiratory syndrome coronavirus 2 (SARS-CoV-2) which causes COVID-19 by serological methods is important to diagnose a current or resolved infection. In this study, we applied a rapid COVID-19 IgM/IgG antibody test and performed serology assessment of antibody response to SARS-CoV-2. In PCR-confirmed COVID-19 patients (n=45), the total antibody detection rate is 92% in hospitalized patients and 79% in non-hospitalized patients. We also studied antibody response in relation to time after symptom onset and disease severity, and observed an increase in antibody reactivity and distinct distribution patterns of IgM and IgG following disease progression. The total IgM and IgG detection is 63% in patients with < 2 weeks from disease onset; 85% in non-hospitalized patients with > 2 weeks disease duration; and 91% in hospitalized patients with > 2 weeks disease duration. We also compared different blood sample types and suggest a potentially higher sensitivity by serum/plasma comparing with whole blood measurement. To study the specificity of the test, we used 69 sera/plasma samples collected between 2016-2018 prior to the COVID-19 pandemic, and obtained a test specificity of 97%. In summary, our study provides a comprehensive validation of the rapid COVID-19 IgM/IgG serology test, and mapped antibody detection patterns in association with disease progress and hospitalization. Our study supports that the rapid COVID-19 IgM/IgG test may be applied to assess the COVID-19 status both at the individual and at a population level.

## INTRODUCTION

The coronavirus disease 2019 (COVID-19) is caused by infection of severe acute respiratory syndrome coronavirus 2 (SARS-CoV-2). As of June 02, 2020, it had caused a total of 6,194,533 cases of infection and 376,320 deaths worldwide.^1^ The COVID-19 pandemic has become a global crisis, and the outbreak has had a substantial impact on the world economy. Resuming from society lockdown to normality depends largely on population screening of COVID-19 immunity, and serology assessment of antibody response to SARS-CoV-2 by blood antibody test is a crucial aid to track the true spread of the pandemic.

In the current clinical practice, the reverse transcription polymerase chain reaction (RT-PCR) is the most commonly used method for diagnosis of COVID-19. The test is performed on upper respiratory tract specimens including nasopharyngeal and throat swabs. Although PCR offers detection of viral RNA, the result may not necessarily reflect the presence of viable virus.^2^ False-negative rates up to 50% may occur due to inappropriate sampling procedure or timing in relation to disease onset.^3,4^ Furthermore, PCR tests are also technically demanding and costly in operation. To compromise the limitation of the PCR test, COVID-19 infection can also be detected by serology assessment of antibody responses to SARS-CoV-2 infection. The antibody test captures the presence of antibodies both during and after the infection and is able to determine if a person has been infected with the virus, even without the development of symptoms. This is particularly important for individuals who present to the health care late after disease onset.^5^ Blood antibody tests may therefore complement the PCR test to accurately diagnose a current or resolved infection, map the extent of community spread of COVID-19, and profile the potential protection from future virus exposure by acquired immunity.

In this study, we assessed antibody responses to SARS-CoV-2 in patients with COVID-19 by a rapid COVID-19 IgM/IgG test. The evaluation was performed on patients from the acute phase of the disease; as well as hospitalized and recovered COVID-19 patients. The specificity of the test was verified on COVID-19 negative blood samples collected prior to the pandemic. We also tested whether different blood sample types (serum/plasma/whole blood) provided the same results for this assay.

## MATERIAL AND METHODS

### STUDY COHORTS AND SAMPLES

#### COVID-19 cohort

Patients with RT-PCR verified COVID-19 were enrolled prospectively either in the convalescence phase (2-8 weeks after start of symptoms) or when they were acutely ill or/and hospitalized. The study was conducted at the Department for Infectious Diseases, Skåne University Hospital, Lund, Sweden. All patients gave their informed consent to participate in the study. Blood was drawn in serum tubes and analyzed directly or after coagulation over night at 8 °C followed by centrifugation at 2 000 g for 10 minutes.

#### Muscle Satellite Cell Study (MSAT) cohort

The MSAT is an ongoing study since 2016 that recruits healthy male volunteers for exercise testing at the Department of Clinical Sciences, Malmö, Lund University, Sweden. All 39 MSAT serum samples included in this analysis were collected between 2016 and 2017.

#### Carotid Plaque Imaging Project (CPIP) cohort

The CPIP is an ongoing cohort since 2006 that recruits patients undergoing carotid endarterectomy at Lund University Hospital to study atherosclerosis and inflammatory and immune markers. Blood samples were collected the day before surgery. For the current study, thirty serum samples collected between 2016 to 2018 were included.

### COVID-19 ANTIBODY TEST IgM/IgG

The serology assessment of COVID-19 antibodies was performed using *ZetaGene COVID-19 Antibody Test IgM/IgG* (ZetaGene, Sweden; www.zetagene.com). The *ZetaGene COVID-19 Antibody Test IgM/IgG* is a lateral flow immunoassay intended for the qualitative detection and differentiation of IgM and IgG antibodies to SARS-CoV-2 in serum, plasma or whole blood samples from patients suspected of COVID-19 infection.

The test cassette consists of: (1) a conjugate pad containing SARS-CoV-2 RBD antigen and rabbit IgG; (2) a nitrocellulose membrane containing IgG line (G) and IgM line (M) coated with anti-human IgG or IgM; and a control line (C) coated with goat anti-rabbit IgG. When the test sample is dispensed into the sample well of the test cassette, the specimen migrates along the cassette. A negative antibody test result is defined as no additional line to the presence of C line. A positive antibody test result is defined as visible G or M line or both in addition to the presence of a C line.

The antibody assessment was conducted according to the manufacturer’s instruction (www.zetagene.com). In brief, 10 μl of serum/plasma or whole blood was dispensed into the test sample well, followed by addition of 100 μl of diluent buffer provided in the kit. The results were visualized between 15-20 minutes.

### ETHICS

The study was performed in accordance with the Declaration of Helsinki. Patients in all study cohorts included gave informed consent to serum donation and study participation.

All studies were approved by the ethical review board: the COVID-19 prospective study (2020-01747); CPIP (472/2005); and MSAT (2015/593).

### STATISTIC ANALYSIS

The data is presented in n (% of total) in respective analysis, and 95% confidence intervals for proportions are calculated according to the Clopper-Pearson method.

## RESULTS

### Qualitative COVID-19 IgM and IgG assessment in response to hospitalization in COVID-19 patients

A total of 45 COVID-19 patients confirmed with RT-PCR were enrolled at the Department for Infectious Diseases, Skåne University Hospital, Lund. In this cohort, twelve patients (27%) had more severe symptoms and were admitted to the hospital; thirty-three patients (73%) had mild symptoms and were home quarantined. Sera from all patients were tested for COVID-19 IgM and IgG reactivity by a rapid COVID-19 antibody test (ZetaGene, Sweden). In 12 hospitalized patients, eleven (92%) were tested antibody positive to SARS-CoV-2, with 9 (75%) positive for both IgM and IgG; two (17%) positive for IgG only (Fig. 1 and 2; Table 1). In 33 non-hospitalized patients, twenty-six (79%) were tested antibody positive, including 15 (45%) IgG positive IgM positive, one (3%) IgM positive only, and 10 (30%) IgG positive only (Fig. 1 and 2; Table 1). Together, the overall positive percentage agreement (PPA) for COVID-19 antibody detection is 92% (95%CI: 62%- 100%) for hospitalized patients, and 79% (95%CI: 61%-91%) for non-hospitalized patients.

**Fig. 1.**
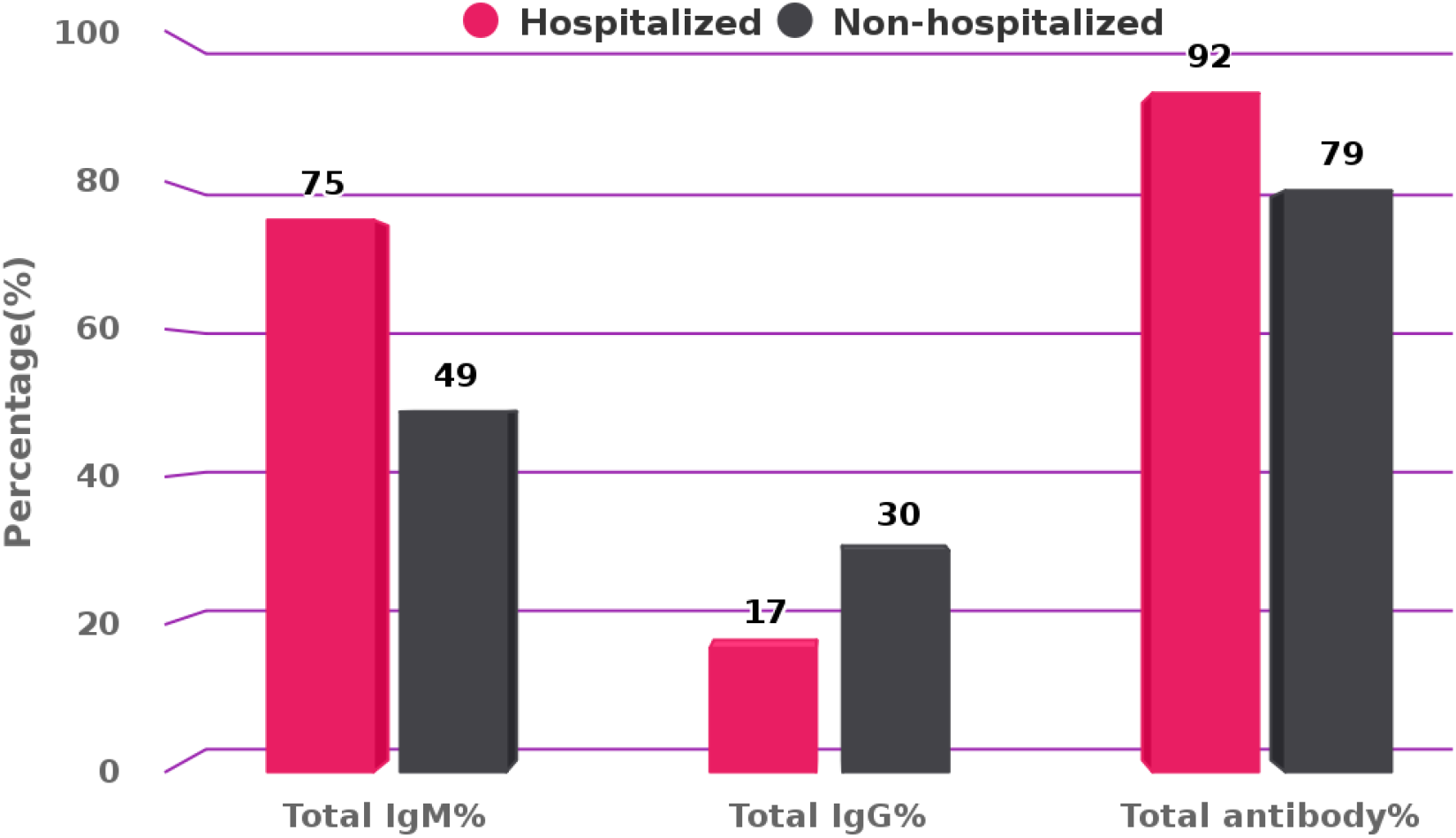
Serum SARS-Cov-2 IgM and IgG antibody detection in response to hospitalization in COVID-19 patients. IgM and IgG reactivity to SARS-CoV-2 was measured in sera in patients with PCR-confirmed COVID-19 (n=45) by lateral flow test *COVID-19 Antibody test IgM/IgG* (ZetaGene, Sweden). Total IgM, IgG and total antibody detection percentage (%) are presented for the hospitalized (red bars, n=12) and nonhospitalized patients (black bars, n=33).

**Fig. 2.**
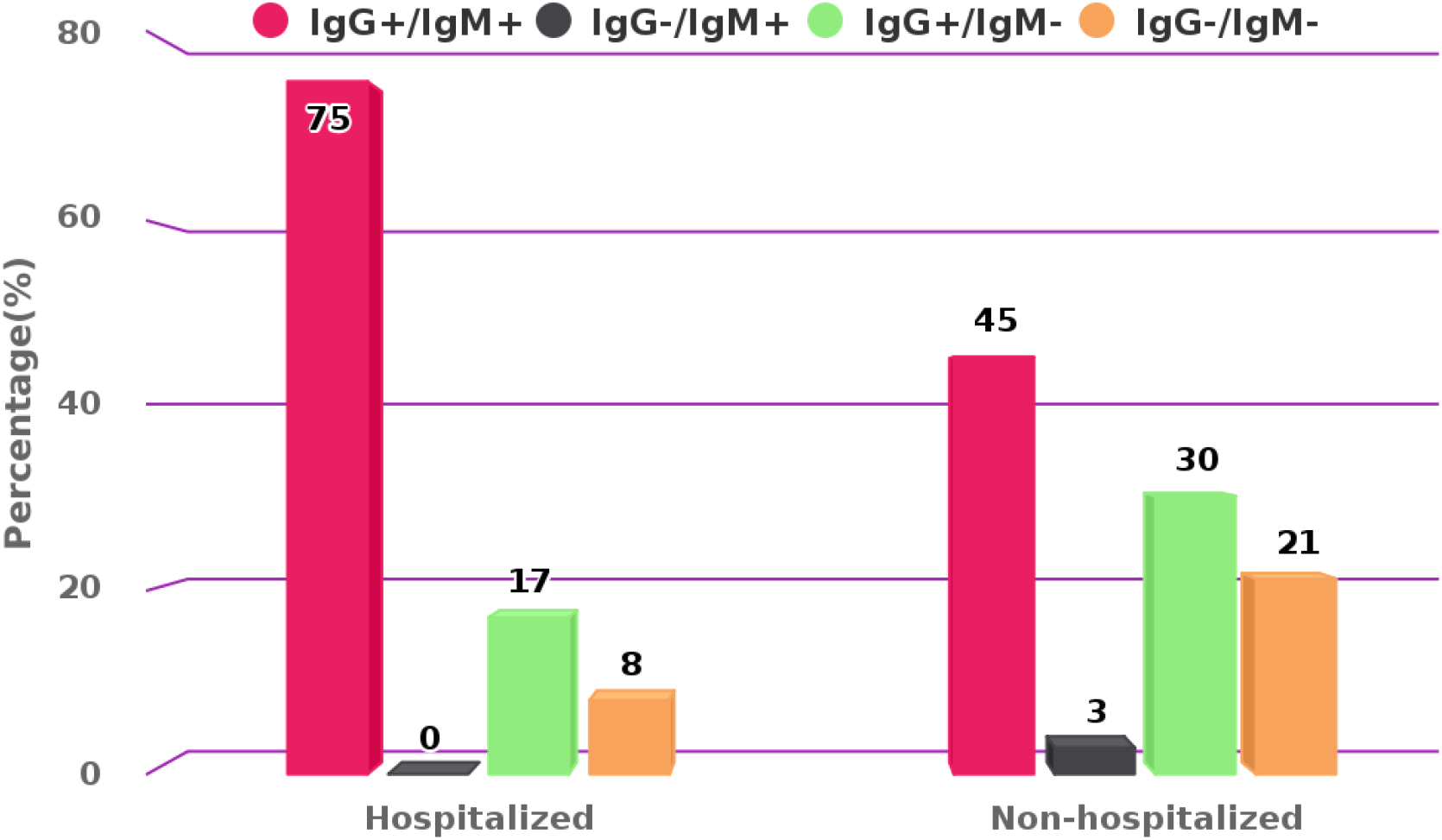
The distribution of COVID-19 IgM and IgG detection in hospitalized *vs*. non-hospitalized patients. Qualitative detection of IgM and IgG to SARS-CoV-2 was assessed in sera of hospitalized (n=12) and non-hospitalized (n=33) COVID-19 patients. IgG and IgM positivity and distribution (%) in the COVID-19 patients are presented in respective color as indicated in the figure.

**Table 1.** Serology assessment of COVID-19 IgG and IgM response by *COVID-19 Antibody Test IgM/IgG* (ZetaGene, Sweden) in sera collected from hospitalized (n=12) and non-hospitalized (n=33) PCR-confirmed COVID-19 patients.

| Hospitalization |  | Hospitalized | Non-hospitalized |
| --- | --- | --- | --- |
| Positive | IgG+/IgM+ | 9 (75%) | 15 (45%) |
|  | IgG-/IgM+ | 0 (0%) | 1 (3%) |
|  | IgG+/IgM- | 2 (17%) | 10 (30%) |
| Negative | IgG-/IgM- | 1 (8%) | 7 (21%) |
| Subtotal |  | 12 (100%) | 33 (100%) |
| Total IgM% |  | 75% | 49% |
| Total IgG% |  | 17% | 30% |
| Total antibody% |  | 92% | 79% |

### Qualitative COVID-19 IgM and IgG assessment in response to time after onset and hospitalization in COVID-19 patients

Previous studies revealed that COVID-19 seroconversion for detectible IgM and IgG begins from the second week of symptom onset.^6^ To better understand the dynamics of IgM/IgG detection in response to time after onset and hospitalization, the cohort patients were grouped into (1) acute patients with < 2 weeks after onset of symptoms (n=8, Table 2); (2) convalescent patients with > 2 weeks after onset, non-hospitalized (n=26, Table 3); (3) convalescent patients with > 2 weeks after onset, hospitalized (n=11, Table 4).

**Table 2.** Serology assessment of COVID-19 IgG and IgM response by *COVID-19 Antibody Test IgM/IgG* (ZetaGene, Sweden) in PCR-confirmed COVID-19 patients with time after onset <2 weeks (n=8).

| <2 weeks after onset |  | PCR-positive | % of Total |
| --- | --- | --- | --- |
| Positive | IgG+/IgM+ | 5 | 63% |
|  | IgG-/IgM+ | 0 | 0% |
|  | IgG+/IgM- | 0 | 0% |
| Negative | IgG-/IgM- | 3 | 37% |
| Subtotal |  | 8 | 100% |
| Total IgM% |  | 5 | 63% |
| Total IgG% |  | 5 | 63% |
| Total antibody% |  | 5 | 63% |

Positive percentage agreement (PPA) = 5/8 (63%, 95%CI: 26%-90%)

**Table 3.**
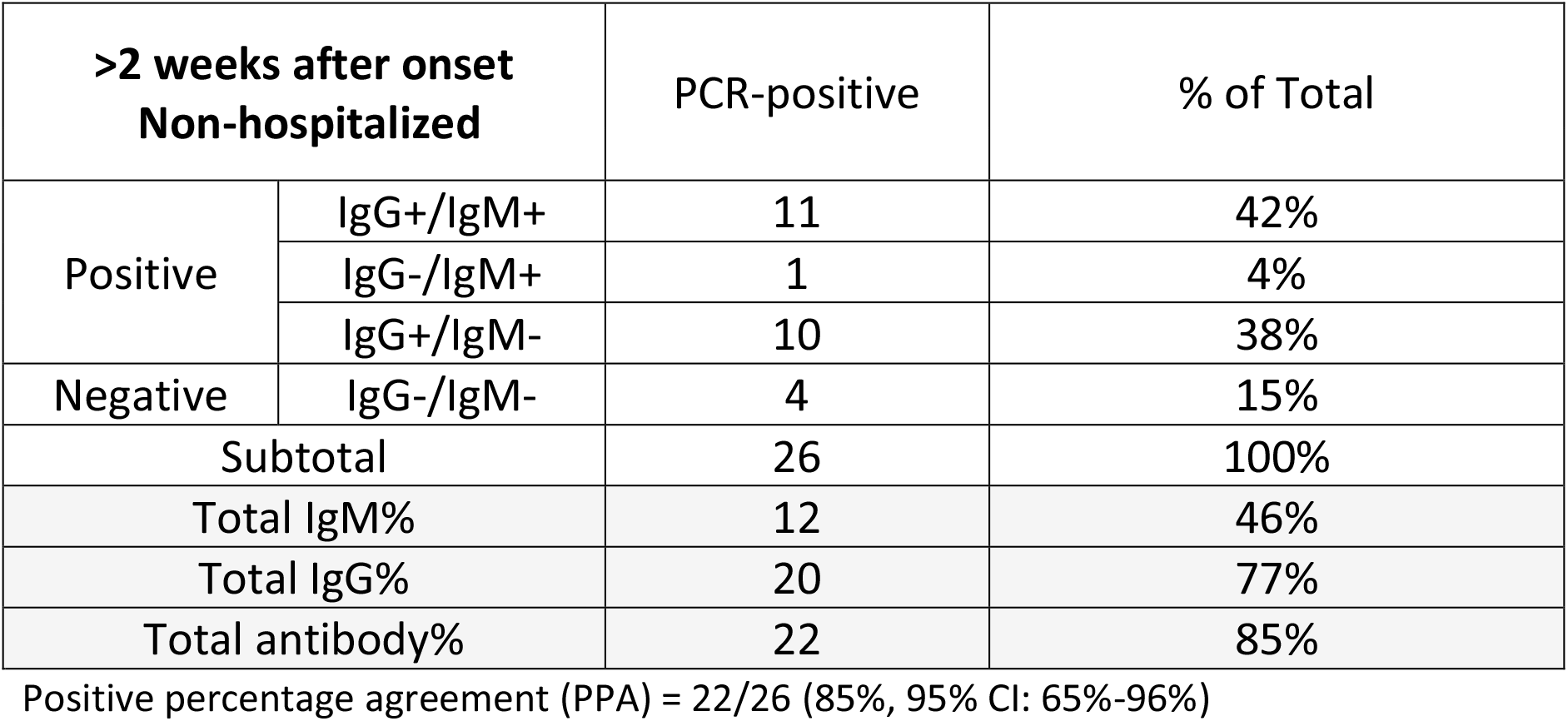
Serology assessment of COVID-19 IgG and IgM response by *COVID-19 Antibody Test IgM/IgG* (ZetaGene, Sweden) in PCR-confirmed non-hospitalized COVID-19 patients with time after onset >2 weeks (n=26).

**Table 4.**
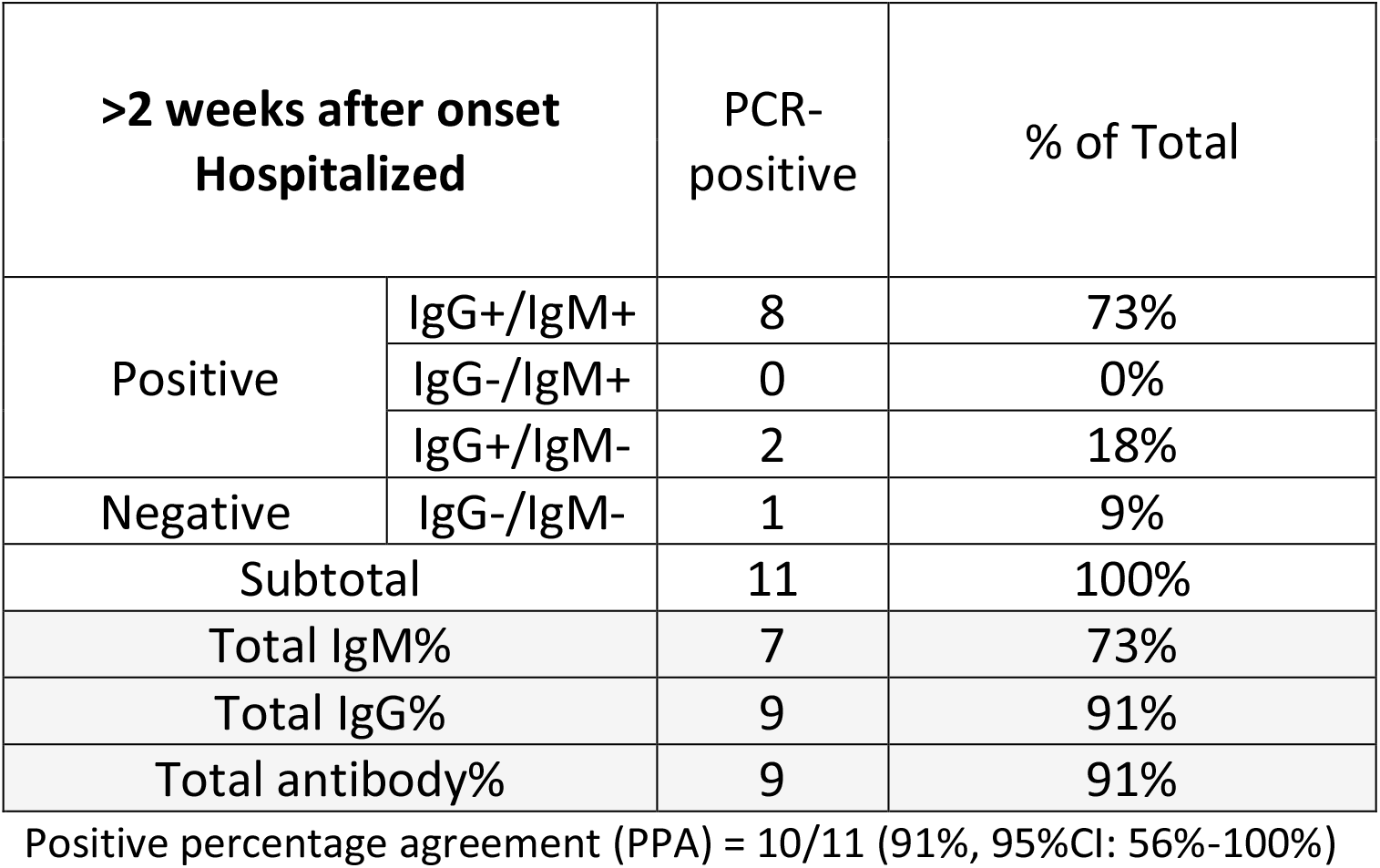
Serology assessment of COVID-19 IgG and IgM response by *COVID-19 Antibody Test IgM/IgG* (ZetaGene, Sweden) in PCR-confirmed hospitalized COVID-19 patients with time after onset >2 weeks (n=11).

| >2 weeks after onset<br>Hospitalized |  | PCR-positive | % of Total |
| --- | --- | --- | --- |
| Positive | IgG+/IgM+ | 8 | 73% |
|  | IgG-/IgM+ | 0 | 0% |
|  | IgG+/IgM- | 2 | 18% |
| Negative | IgG-/IgM- | 1 | 9% |
| Subtotal |  | 11 | 100% |
| Total IgM% |  | 7 | 73% |
| Total IgG% |  | 9 | 91% |
| Total antibody% |  | 9 | 91% |

Positive percentage agreement (PPA) = 10/11 (91%, 95%CI: 56%-100%)

In sera from 8 acute patients (7 non-hospitalized and 1 hospitalized) with less than 2 weeks after onset, five patients were tested IgG and IgM positive (63%), and 3 were negative to IgG or IgM (37%, Fig. 3 and 4, Table 2). The overall PPA for COVID-19 antibody detection in this subgroup is 63% (95%CI: 26%-90%).

**Fig. 3.**
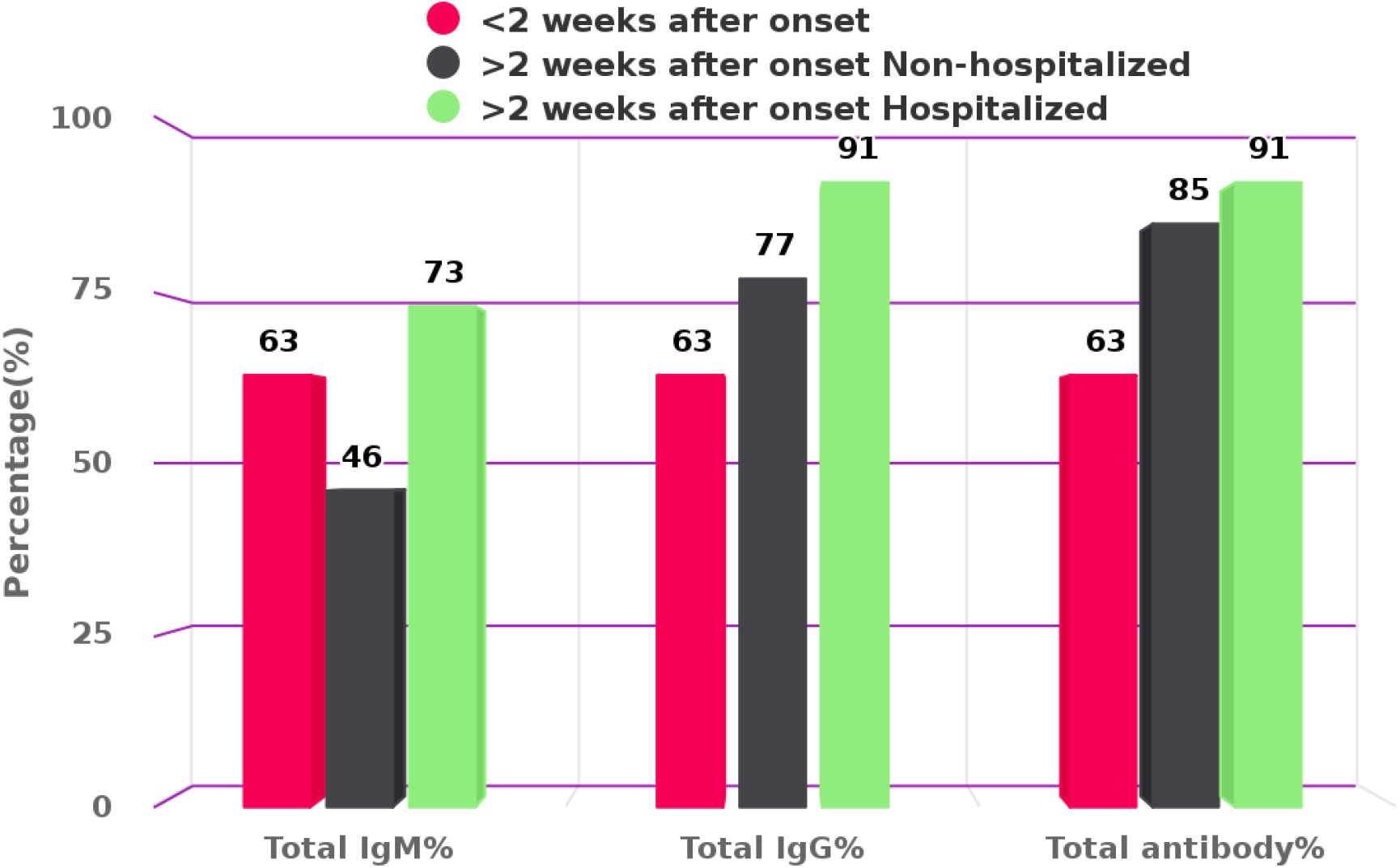
The distribution of antibody detection in response to time from onset and hospitalization. Total IgM, IgG and total antibody detection percentage (%) are presented for PCR-confirmed COVID-19 patients divided into the following groups: (1) acute patients with < 2 weeks (w) after onset (n=8); (2) convalescent patients with > 2 weeks after onset, non-hospitalized (n=26); (3) convalescent patients with > 2 weeks after onset, hospitalized (n=11). Different groups are presented in respective colors as indicated in the figure.

**Fig. 4.**
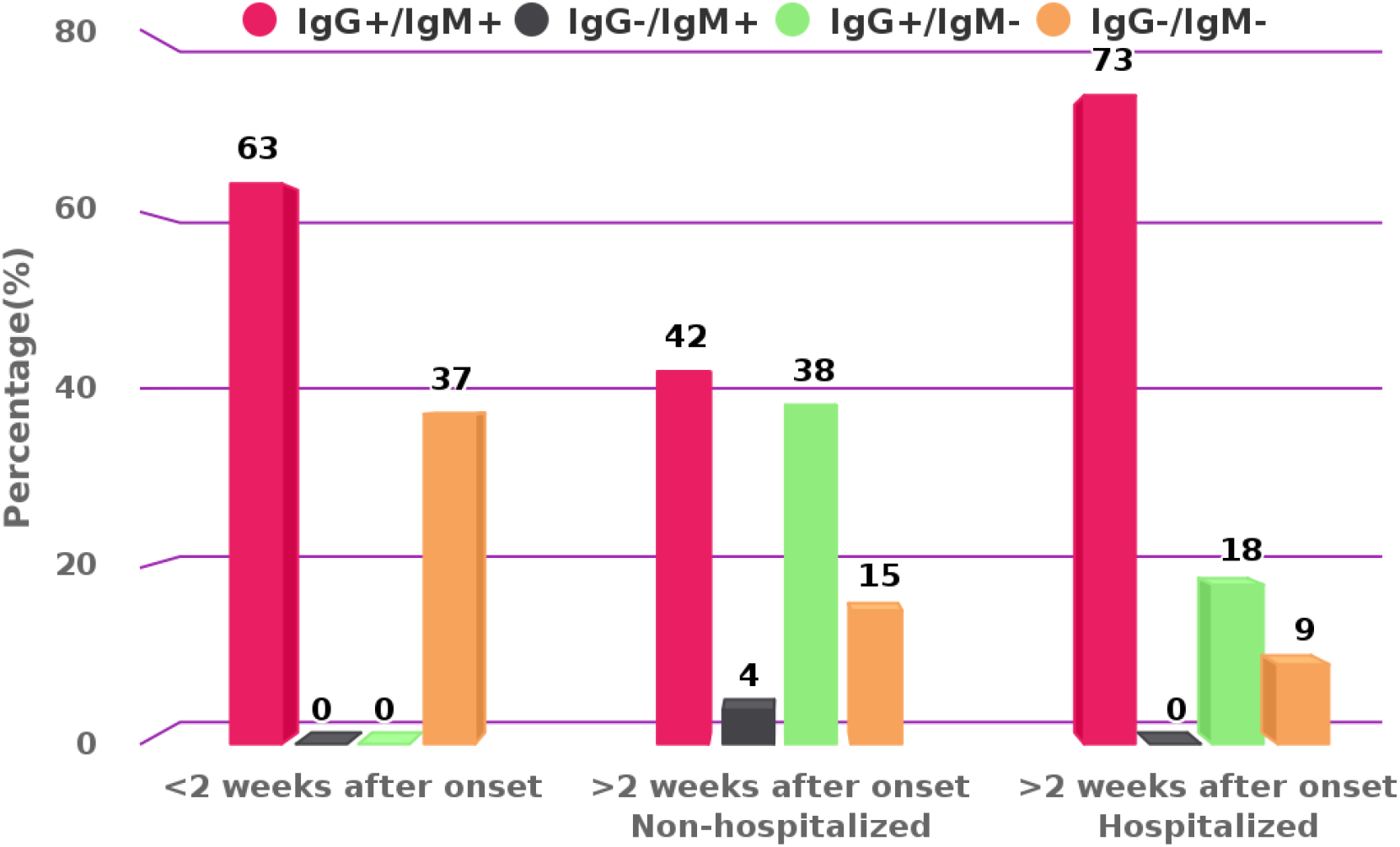
The dynamics of COVID-19 IgM/IgG detection in response to time from onset and hospitalization. Qualitative detection of IgM and IgG to SARS-CoV-2 is mapped in COVID-19 cohort patients from the following groups: (1) acute patients with < 2 weeks (w) after onset (n=8); (2) convalescent patients with > 2 weeks after onset, non-hospitalized (n=26); (3) convalescent patients with > 2 weeks after onset, hospitalized (n=11). IgM and IgG detection percentage (%) is presented in respective color as indicated in the figure.

In convalescent sera from 26 non-hospitalized patients with > 2 weeks after onset, eleven (42%) were positive for both IgG and IgM, one (4%) was only IgM positive, ten (38%) were only IgG positive, and 4 (15%) were IgG/IgM negative (Fig. 3 and 4, Table 4). The overall PPA for COVDI-19 antibody detection was 85% (95%CI: 65%-96%) for this subgroup.

In convalescent sera from 11 hospitalized patients with > 2 weeks after onset, seven (73%) patients were positive for both IgG and IgM, and 2 (18%) were IgG positive only. There was also 1 (9%) patient with no detectible IgG or IgM (Fig. 3 and 4, Table 4). The overall PPA in this subgroup for COVDI-19 antibody detection was 91% (95%CI: 56%- 100%).

The distribution of antibody detection in the above three groups of patients is 63%, 46% and 73% for total IgM; 63%, 77% and 91% for total IgG; and 63%, 85% and 91% for total antibody (Fig. 3).

Among all individuals, three patients (ID 2, 5 and 12) were tested at both < 2 weeks and > 2 weeks. Patients 2 and 5 were hospitalized at both samplings, and patient 12 was home quarantined. Patient 2 was IgG and IgM positive at first sampling, and converted to IgG positive IgM negative at the second sampling. Patient 5 was IgG and IgM positive at both time points. Patient 12 was antibody negative at < 2 weeks, and converted to IgG and IgM positive at > 2 weeks.

### Comparison of antibody detection between whole blood, serum and plasma sampling

Serum and plasma antibody detection from the same individuals were compared in 8 patients, and no difference between the two measurements was observed. We also compared serum and whole blood readout from 19 patients. In 18 patients, serum and whole blood gave uniform antibody detection. However, in 1 patient, we observed that IgG positive was only detected in the serum but not in the whole blood.

### Verification of IgM and IgG reactivity in pre-COVID-19 control cohorts

For accurate interpretation of the rapid IgM/IgG serology assessment of COVID-19 sera, and to determine the specificity of this rapid test, we performed qualitative measurements of IgM and IgG in two control cohorts with blood samples collected prior to the COVID-19 pandemic (2016-2018).

#### A healthy control cohort - MSAT cohort

The MSAT cohort includes healthy, well-trained males recruited for exercise tests conducted at the Department of Clinical Sciences, Malmö, Lund University, with the characteristics of age 37.4 ± 8.3 years [mean ± SD], BMI 24.4 ± 2.4 kg/m^2^ [mean ± SD], and VO2MAX = 52.0 ± 8.1 ml/kg/min [mean ± SD]. Thirty-nine serum samples collected between 2016-2017 were tested for COVID-19 IgM/IgG reactivity. Of 39 MSAT samples, thirty-eight were tested negative for IgM or IgG, and 1 was tested weak IgG positive. The overall negative percentage agreement (NPA) for negative antibody detection is 97% (95%CI: 87%-100%).

#### A cardiovascular disease (CVD) control cohort - CPIP cohort

The CPIP cohort has continuous recruitment of patients since 2006 with advanced atherosclerosis and confirmed CVD at the Department of Clinical Sciences, Lund University. Thirty serum samples were randomly selected from the CPIP cohort, which include 10 samples from 2016, 10 from 2017 and 10 from 2018. The characteristics of the individuals included in this study are as follows: age 71 ± 8 years [mean ± SD], BMI 27 ± 4 [mean ± SD], 21 males and 9 females. Of the 30 samples, twenty-nine (97%) were tested negative for IgM or IgG, and 1 sample showed very weak IgM positive. The overall NPA for negative antibody detection is 97% (95%CI: 83%-100%).

By combining both MSAT and CPIP cohorts, we obtained an overall NPA 97% (95%CI: 90%-100%) for total negative antibody detection.

## DISCUSSION

In the global response to fight the COVID-19 pandemic, it remains crucial to profile the true population epidemic, and provide knowledge on the nature of protection by acquired active immunity. This is especially important in preparation for lifting confinement restrictions and preventing potential new waves of SARS-CoV-2 spread. Serological testing of COVID-19 antibody response is therefore a critical component in overcoming the pandemic. In this study, we performed detailed serology assessment on SARS-CoV-2 antibody response in patients with COVID-19 by a rapid IgM/IgG test. We compared different blood sample types and evaluated antibody responses in relation to the time from onset and hospitalization. Furthermore, we verified test specificity in control cohorts recruited prior to the COVID-19 pandemic.

Recent studies reveal that the spike glycoprotein (S) of SARS-CoV-2 mediates receptor binding on host cells, membrane fusion and virus entry. Additionally, it is the main target for neutralizing antibodies.^7^ Each monomer of trimeric S protein contains two subunits, S1 and S2, mediating attachment and membrane fusion, respectively, with the S1 region being the major immune-epitope for antibody binding. In this study, we employed a lateral flow test that detects antibodies binding to the receptor-binding domain (RBD) of S1 protein. The test offers qualitative assessment and indicates the presence or absence of SARS-CoV-2 antibodies. The test has been previously evaluated on 38 COVID-19 hospitalized patients and 228 healthy controls during Wuhan COVID-19 outbreak in China, and achieved a sensitivity and a specificity of 95% and 97% (www.zetagene.com). In the current study, we observed that 92% of hospitalized patients have detectable antibodies, which is similar to the clinical validation in China (unpublished). The negative test results may be explained by a combination of test sensitivity, the complexity of antibody development biology, as well as the heterogeneity in antibody titres among individuals.

A previous study revealed that IgM kinetics is notably different between COVID-19 ICU and non-ICU patients. In patients with mild symptoms, IgM levels start to decline after 2 weeks while IgG continues to increase, while in patients with severe symptoms, IgM levels remains static or increases even after 4 weeks.^9^ In our study, IgM was detected in 49% of non-hospitalized convalescent patients, and 75% in hospitalized patients (Fig. 1). This observation suggest that IgM may be present in the blood for a longer period in hospitalized patients. Furthermore, it has been shown that high titers of IgG antibodies positively correlate with neutralizing antibodies.^10^ In our study we also observed higher prevalence of IgG detection in non-hospitalized vs. hospitalized convalescent patients (30% vs. 17%, Fig. 1). Furthermore, we mapped a clear increase in the antibody response from early disease onset (< 2 weeks, 63%) to a later time point (>2 weeks, 85% and 91%, Fig. 3 and 4), which is in line with the antibody development profile following COVID-19 progression and an earlier investigation in Sweden.^8^

A previous study on COVID-19 recovered patients showed that there is a heterogeneity in antibody titers among the recovered patients, and around 15% of recovered patients had very low antibody titers or even levels below the threshold of detection.^11^ This observation is also in line with our study on recovered patients, where we observed that 85% of non-hospitalized recovered patients and 91% of hospitalized recovered patients had detectable COVID-19 antibodies (Fig. 3).

We compared antibody detection in whole blood, serum and plasma. In one patient out of 19 patients, IgG positivity was only detected in serum but not in whole blood. Also, upon comparison of the sensitivity in serum versus plasma, we did not observe any difference in the 8 patients where both plasma and serum were analyzed. Albeit suggestive, clinical assessment of COVID-19 antibody reactivity performed on serum/plasma is likely to be more accurate than that of whole blood. Further evaluation is needed to compare pinprick blood, whole blood and serum/plasma sensitivity.

The choice of negative control cohorts also warrants explanation. The COVID-19 pandemic was reported to have reached Sweden on 31 January 2020 with the first clinically confirmed case.^12^ The epidemic outbreak of COVID-19 in Sweden started on 26 February, with confirmed community transmission on 9 March in the Stockholm region. However, according to the Public Health Agency of Sweden, it is likely that there were individual cases of COVID-19 in Sweden as early as November 2019. Therefore, we chose to use cohorts with blood samples collected before 2019 as the negative controls of the test. We tested both a healthy cohort and a diseased cohort with cardiovascular morbidity, and both cohorts displayed uniform specificity.

In summary, our study provides a comprehensive validation of the rapid COVID-19 IgM/IgG serology assessment. Also, the antibody detection patterns in association with disease progress and hospitalization was mapped, which provides a potential reference for accurate clinical antibody assessment interpretation. The rapid COVID-19 IgM/IgG test may be a valuable application for COVID-19 clinical diagnosis, and can be applied as a powerful tool to assess the COVID-19 status at both individual and population level.

## Data Availability

The authors confirm that the data supporting the findings of this study are available within the article.

## ACKNOWLEDGMENTS

This study was supported by the Swedish Research Council, Strategic Research Area Exodiab, Dnr 2009-1039; and the Swedish Foundation for Strategic Research Dnr IRC15- 0067, the Swedish Research Council (OH), the Crafoord foundation (OH), Governmental funding of clinical research within the NHS (National Health Services) (to MR, AB and OH), the Novo Nordisk foundation (OH) and the Påhlsson foundation (OH). The CPIP study was supported by grants to IG from the Swedish Research Council, Swedish Heart and Lung Foundation, Skåne University Hospital, Stroke Foundation, ALF grants Region Skåne.

We thank Esa Laurila and Ylva Wessman (at Lund University) for technical assistance with the MSAT study, Nils Fernström for help with COVID-19 patient serum collection, Andreas Edsfeldt, Ana Persson, Mihaela Nitulescu for support with CPIP sample collection.

The *COVID-19 Antibody Test IgM/IgG* kits were donated by ZetaGene Ltd. (Sweden).

## DECLARATION OF INTERESTS

TS, PB, AB, RY, JS, OE, KFE, OH, LG, IG and MR declare no conflict of interest. YDM is the founder of and has an equity interest in ZetaGene Ltd. (Sweden), a company that is developing microfluidic technologies for point-of-care diagnostic solutions.

## NOTES ON CONTRIBUTORS

***Yang De Marinis*** is Associate Professor at the Department of Clinical Sciences, Lund University, Sweden. She received her PhD on electrophysiology at Lund University, and is currently a principle investigator with focus on chromatin immunoprecipitation sequencing, genome editing, clinical cohort studies and machine learning application. She also holds a visiting chair at the Clinical Research Hospital, Chinese Academy of Sciences; the Division of Life Sciences of Medicine, University of Science and Technology of China. She is currently involved in several projects concerning COVID-19.

***Magnus Rasmussen*** is Professor of infection medicine and consultant in infectious diseases. He is an expert on bacterial endocarditis and is currently involved in several projects concerning COVID-19.

***Torgny Sunnerhagen*** is a licensed physician and PhD student of infection medicine. His areas of expertise are bacterial endocarditis and diagnostic risk classification systems. At present he is involved in projects regarding COVID-19.

***Pradeep Bompada*** is PhD student at Department of Clinical Sciences, Lund University, Sweden. He received his Master’s degree in molecular biology from Lund University. He is currently investigating epigenetic mechanisms and blood-based epigenetic biomarkers associated with type 2 diabetes.

***Anna Bläckberg*** is a resident physician in infectious diseases and PhD student of infection medicine. Her areas of expertise are endocarditis and *Streptococcus* bacteremia. At present she is involved and working with projects regarding COVID-19.

***Runtao Yang*** is currently a lecturer at the School of Mechanical, Electrical and Information Engineering, Shandong University at Weihai, China. His research areas include machine learning, bioinformatics, and system biology.

***Joel Svensson*** is a M.D. and specialist in Clinical Chemistry at Skåne University hospital, Malmö, Sweden. In his PhD project at the Department of Laboratory Medicine, Lund University, tick-borne pathogens are investigated among Swedish populations, especially the hemoparasite Babesia and the Tick-Borne Encephalitis Virus.

***Ola Ekström*** is a Medical Doctor and PhD student with a special interest in exercise metabolism and diabetes. He is a research physician with experience from both sponsor-and investigator initiated clinical trials.

***Karl-Fredrik Eriksson*** is Associate Professor at the Department of Clinical Sciences. His research focuses on vascular diseases and related clinical studies.

***Ola Hansson*** is since 2013 Associate Professor of functional genomics, currently heading the research unit Genomics, Diabetes and Endocrinology at the Department of Clinical Sciences, Lund university, Sweden. Since 2018 he holds a researcher position at the Institute for Molecular Medicine Finland (FIMM), Helsinki University, Finland. The main research interest is to understand how genetic variation influences skeletal muscle function and whole-body metabolism - implications primarily for human health, but also for fundamental evolutionary questions. The overarching goal is to tailor, for the individual, the most beneficial exercise program to counteract genetic predisposition to metabolic disease and type 2 diabetes.

***Isabel Goncalves*** is professor and senior consultant of Cardiology, Skåne University Hospital, Lund University. Her areas of expertise are atherosclerosis, its underlying mechanisms (particular in inflammation and immunity), as well as development of tools to identify patients at risk for heart attacks and strokes.

*Leif Groop*, **M.D., Ph.D. is currently Senior Professor at Lund University, Sweden and Research Director at Institute of Molecular Medicine Finland (FIMM) Helsinki, Finland. He was in 1993 elected Professor in Endocrinology at Lund University and has served as Chief Physician of the Endocrine Clinic as well Director of Lund University Diabetes Centre (LUDC). He is a member of the Swedish Royal Academy of Science, Distinguished Visiting Professor at University of Science and Technology of China**.

## Notes

### Author Declarations

ETHICS The study was performed in accordance with the Declaration of Helsinki. Patients in all study cohorts included gave informed consent to serum donation and study participation. All studies were approved by the ethical review board: the COVID-19 prospective study (2020-01747); CPIP (472/2005); and MSAT (2015/593).

